# The Effect of Ultraviolet C Radiation Against SARS-CoV-2 Inoculated N95 Respirators

**DOI:** 10.1101/2020.05.31.20118588

**Authors:** David M. Ozog, Jonathan Z. Sexton, Shanthi Narla, Carla D. Pretto-Kernahan, Carmen Mirabelli, Henry W. Lim, Iltefat H. Hamzavi, Robert J. Tibbetts, Qing-Sheng Mi

## Abstract

Since March 31^st^, 2020, during the height of the pandemic, we have decontaminated thousands of 3M 1860 respirators with Ultraviolet C (UVC) for our frontline workers. There is no published peer-reviewed data regarding the dose required to effectively disinfect SARS-CoV-2 on N95 filtering facepiece respirators (FFRs). Four different locations (facepiece and strap) on 5 different N95 FFR models (3M 1860, 8210, 8511, 9211; Moldex 1511) were inoculated with a 10 μL drop of SARS-CoV-2 viral stock (8 × 10^7^ TCID_50_/mL). The outside-facing and wearer-facing surfaces of the respirators were each irradiated with a dose of 1.5 J/cm^2^ UVC (254 nm).

Viable SARS-CoV-2 was quantified by a median tissue culture infectious dose assay (TCID_50_). UVC delivered using a dose of 1.5 J/cm^2^, to each side, was an effective method of decontamination for the facepieces of 3M 1860 and Moldex 1511, and for the straps of 3M 8210 and the Moldex 1511. This dose is an appropriate decontamination method to facilitate reuse of respirators for healthcare personnel when applied to certain models/materials. Increasing the dose may improve decontamination for the other models and straps; however, UVC radiation can degrade certain polymers in a dose dependent manner, and the effects may vary greatly between different models. Therefore, fit-testing of UVC decontaminated respirators must be performed each time a new model and/or dose is introduced into the healthcare system.

## Introduction

The shortage of personal protective equipment (PPE) is affecting healthcare workers worldwide during the coronavirus disease 2019 (COVID-19) pandemic. The ability to decontaminate and reuse N95 filtering facepiece respirators (FFRs) is a partial solution to the current shortage.(1) We recently proposed decontaminating respirators with repurposed dermatology office phototherapy devices, which serve as a platform for ultraviolet C (UVC) germicidal disinfection.(2) On March 31^st^, during the height of the pandemic, Henry Ford Health System (HFHS) began decontaminating 3M 1860 respirators with UVC and returning them to their original users. Since then, several thousand respirators have been decontaminated.

Previous studies have shown that UVC can inactivate other coronaviruses including Severe Acute Respiratory Syndrome coronavirus (SARS-CoV) and Middle East Respiratory Syndrome coronavirus (MERS-CoV).(3, 4) However, there is no peer-reviewed published data showing the effective disinfection of the causative agent of COVID-19, SARS-CoV-2 by UVC, on intact N95 filtering facepiece respirators (FFRs). Consequently, this was causing significant anxiety in our frontline workers using decontaminated PPE.

The objective of this study was to determine the effect of UVC on decontamination of SARS-CoV-2-innoculated N95 respirators using a variety of FFRs that are available to healthcare employees at Henry Ford Health System (HFHS) in Detroit, MI.

## Methods

The study was performed as a collaboration between the HFHS and the University of Michigan. All study procedures were approved and conducted according to the University of Michigan Institutional Biosafety Committee BSL3 (Biosafety Level 3). The appropriate training and medical surveillance for experimental procedures and manipulations performed in the BSL3 facility were satisfied by all individuals directly involved in laboratory testing at University of Michigan.

### Virus and Preparation of Viral Stocks

The SARS-CoV-2 strain used was USA-WA1/2020 NR-52281. Viral stocks of SARS-COV-2 were obtained from the Biodefense and Emerging Infections Research Resources Repository and were propagated in Vero-E6 cells grown in Dulbecco’s Modified Eagle Medium (DMEM) without phenol red, with 2% Fetal Bovine Serum (FBS), L-glutamine, penicillin/streptomycin, non-essential amino acids, and hydroxyethyl piperazineethanesulfonic acid (HEPES). The virus stock was purposely produced in a phenol red-free medium to avoid photodegradation or photooxidation that may affect the results. For stock virus titration, aliquots of viral stock were applied on confluent Vero-E6 cells in 96-well plates for a 50% tissue culture infectious dose (TCID_50_) assay. Viral stocks were determined to be 8 × 10^7^ TCID_50_/mL.

### Test respirators and UVGI device

Respirators were tested 100% intact and included the following models: 3M 1860 (St. Paul, MN); 3M 8210 (St. Paul, MN); Moldex 1511 (Culver City, CA); 3M 8511 (St. Paul, MN); and 3M 9211(St. Paul, MN).The low-pressure mercury lamp ultraviolet germicidal irradiation device (UVGI) (254 nm, 1 series) was manufactured by Daavlin (Byron, OH), with custom dimensions (22 in. × 10 in. × 8 in) to fit under the BSL3 biosafety hood. The irradiance of the device was approximately 16.5 mW/cm^2^ at a distance of 11.5 cm from the lamps (approximately at the apex of the N95 respirator). The UVGI device used 4 lamps, spaced 4.5 cm apart. In comparison, the devices used by HFHS to decontaminate respirators for healthcare personnel had an irradiance of approximately 10 mW/cm^2^ at a distance of 11.5 cm from the lamps. This UVGI device had 10 lamps, spaced 11 cm apart. Despite the differences, the units are similar in performance. Prior to initiating the experiment, the irradiance of the device was measured, and a built-in dosimeter was calibrated to adjust the irradiation.

### Decontamination studies

Intact FFRs in a donned position were inoculated on the outside-facing surface with a single 10 μL drop of viral stock (8 × 10^7^ TCID_50_/mL) on 4 areas to account for differing received doses on complex surfaces: nosepiece, apex, chin-piece, and strap **(Figure 1A)**. Inoculated respirators were dried in a biosafety cabinet at room temperature for 40 minutes. For each N95 respirator model, FFRs were UVC-irradiated or left untreated as positive controls for viral load recovery. The respirators were then placed under the UVGI device, in the center, and were individually treated with a dose of 1.5 J/cm^2^. Then, they were rotated and the wearer-facing side of the N95 was again irradiated with 1.5 J/cm^2^. The irradiation time for each side was approximately 60-70 seconds. The device does not generate any heat; as such, all FFRs were exposed to UVC at room temperature. Immediately after the completion of the irradiation, 4 mm circles containing the inoculated surface were obtained with a leather belt eyelet hole punch tool and were placed in 300 μL (microliters) of PBS for 1 hour at room temperature.

**Figure 1.**
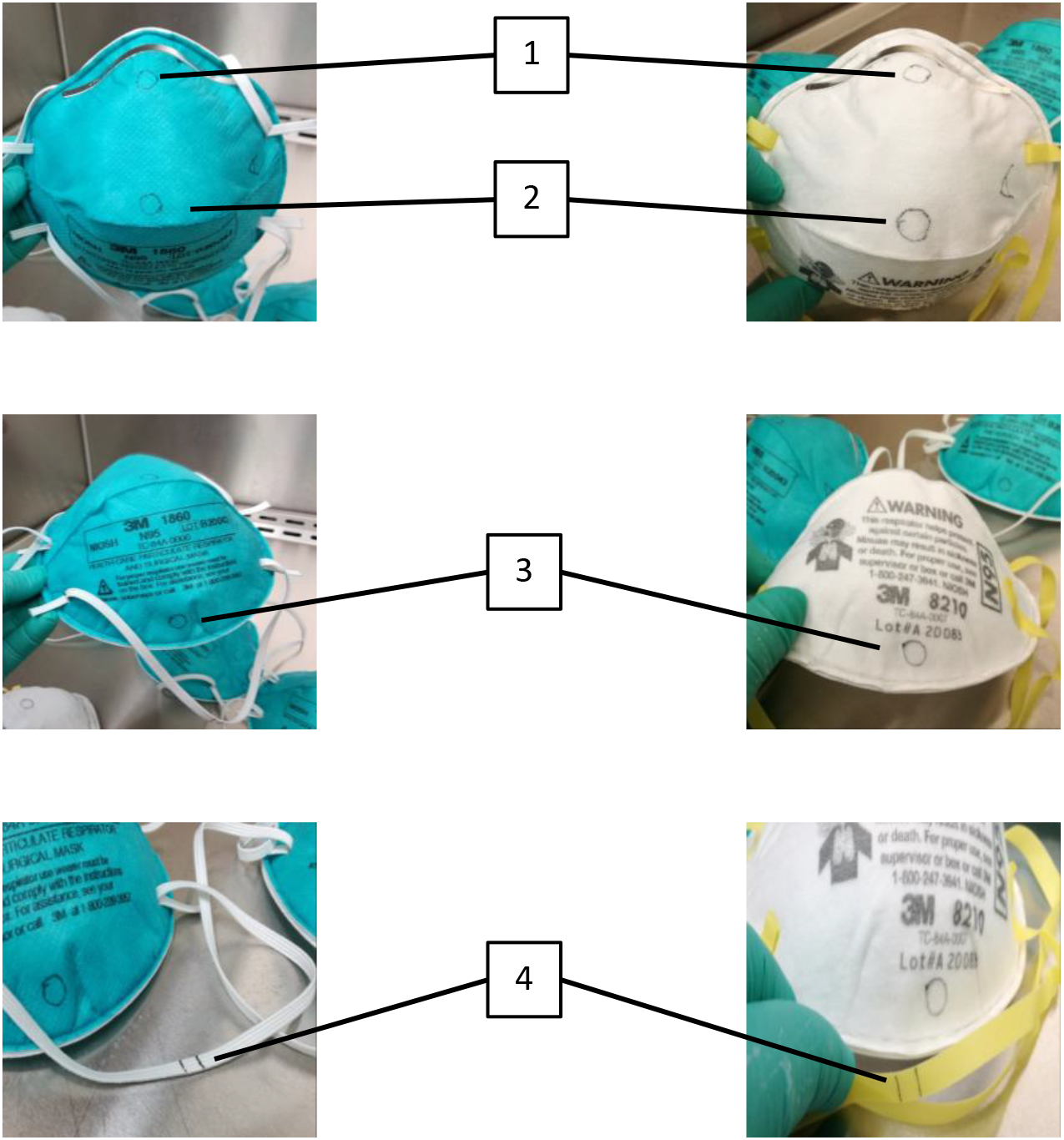
Locations 1-4 (Nosepiece, Apex, Chin, Strap) on models 1860 and 8210. *Simimilar locations were sampled on each of the five N95 respirators.

Recovered viral loads were determined by TCID_50_ assay of the absorbed samples. Briefly, 25 μL aliquots of serially 10-fold diluted samples were inoculated into 96-well plates with a Vero-E6 cell monolayer in sextuplicate and cultured in DMEM with 2% FBS, L-glutamine, penicillin/streptomycin, non-essential amino acids, and HEPES. The plates were observed for cytopathic effects for 4 days. Viral titer was calculated with the Reed and Müench endpoint method.(5) Viral yields were expressed as total TCID_50_ recovered in 300 μL or TCID_50_/4mm punch. TCID_50_ negative controls were cells with media only and were included on each plate assayed. All negative controls had no cytopathic effect (CPE). The limit of detection (LOD) for the TCID_50_ assay was determined to be 10^1.3^ TCID_50_ / 4 mm punch. If the amount of viral particles was below the LOD, then a theoretical, yet low content of viruses may be present.(6) However, an absence of CPE in the Vero E6 cells at 4 days post inoculation indicates a loss of infectivity and is evidence of inactivation of the SARS-CoV-2 samples **(Figure 1B)**.(6) We considered effective decontamination to be results below the LOD with no CPE, the elimination of all infectious SARS-CoV-2.

**Figure 1B.**
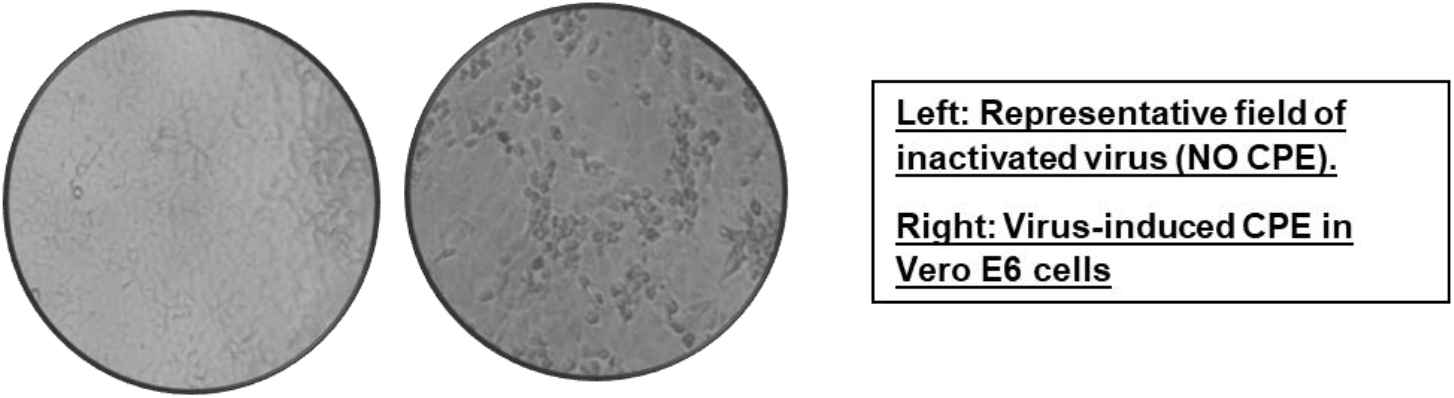
Bright-field microscopy of wells with Vero-E6 cells and SARS-CoV-2. *CPE = cytopathic effect

## Results

Following preliminary testing **(Appendix Figure 1, Appendix Table 1)**, virus inoculation was performed on all 5 types of respirators. For each type, three were irradiated with UVC, and one was not irradiated to serve as positive control. Similar results were seen for the 3M 1860 respirators as in the preliminary study. All facepiece locations were below the LOD with absent CPE. Two straps were above the LOD, and one strap was below the LOD with absent CPE. Sufficient virus (≤ 1 log reduction) was recovered from the untreated positive controls on all facepiece locations; however, a lower yield was recovered from the untreated control strap **(Figure 2, Appendix Table 2)**.

**Figure 2.**
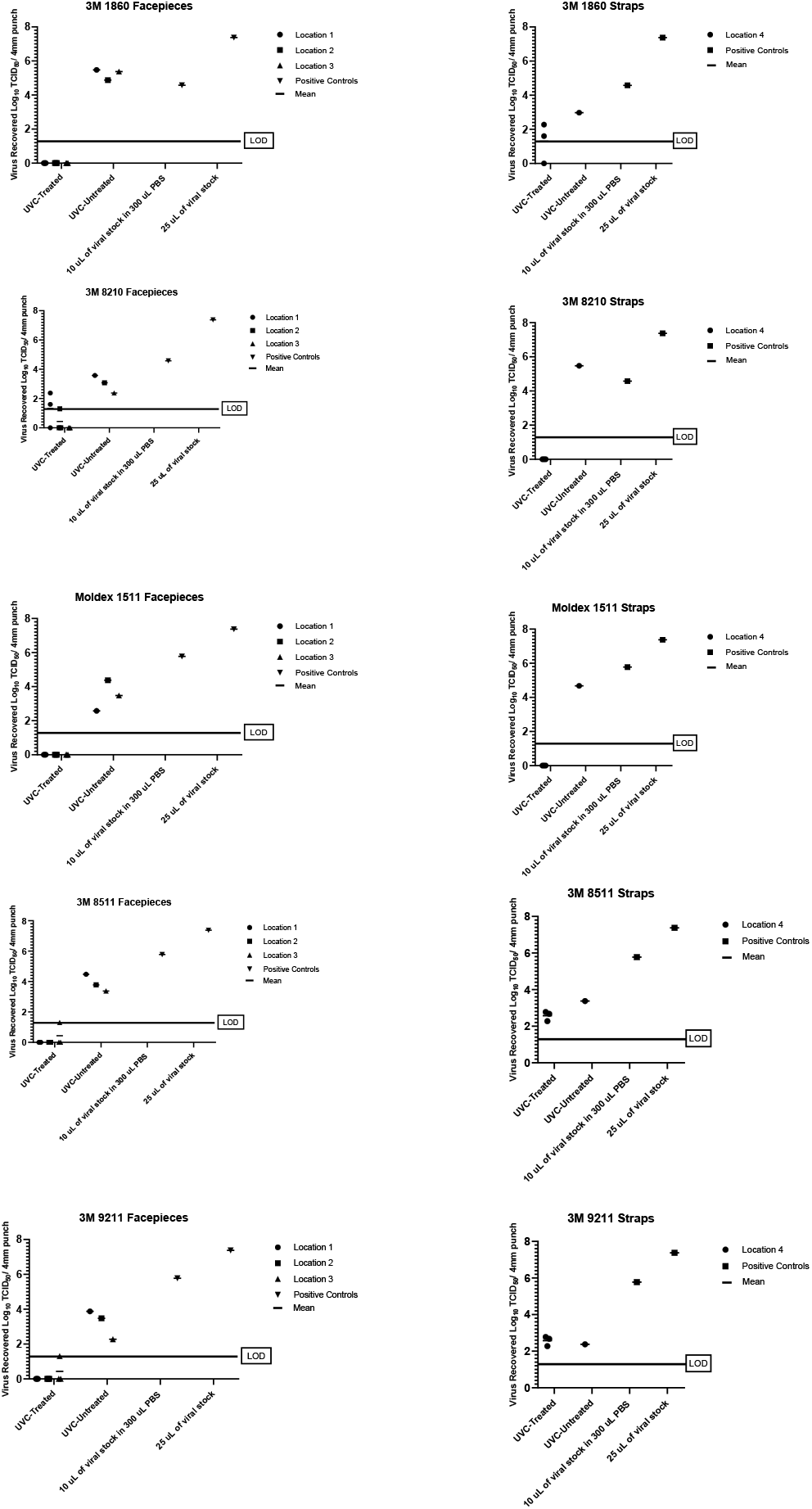
Recovered SARS-CoV-2 TCID50 / 4mm punch. *Wells that were below the limit of detection (LOD) and had no cytopathic effect were arbitrarily assigned the value of zero to represent this phenomenon in the above graphs.

On the 3M 8210 respirators, location 1 had two respirators above the LOD, and one respirator below the LOD with absent CPE. Location 2 had one FFR at the LOD, and two FFRs below the LOD with absent CPE. Location 3 and all the straps were below the LOD with absent CPE. Lower virus yields were recovered from the untreated positive control on all facepiece locations. In contrast, the strap did not absorb the droplet and a sufficient yield was obtained. **(Figure 2, Appendix Table 2)**. Of note, the amount of virus recovered from the strap on the untreated positive control was higher than the untreated control virus stock (10 μL in PBS control). This could have been due to a loss of viral titer of the stock as the control sat in PBS solution for an hour during the experiment.

On the Moldex 1511, all facepiece locations and straps were below the LOD with absent CPE. However, there was a lower virus recovery from certain facepiece locations (1 and 3) on the untreated positive respirator **(Figure 2, Appendix Table 2)**.

For the 3M 8511 and 3M 9211, locations 1 and 2 had FFRs all below the LOD with absent CPE. Location 3 had one FFR at the LOD, and two respirators below the LOD with absent CPE. All the straps were above the LOD. All facepiece locations and the strap on the untreated control had lower virus recovery as compared to the 10 μL in PBS control **(Figure 2, Appendix Table 2)**.

## Discussion

Five N95 respirator models were inoculated with SARS-CoV-2 and tested. UVC delivered using a dose of 1.5 J/cm^2^, to each side, was an effective method of decontamination for the facepieces of 3M 1860 and Moldex 1511, and for the straps of 3M 8210 and the Moldex 1511. This is consistent with previous results using H1N1 influenza demonstrating that UVC decontamination is dependent on model and material type. Mills et al(7) and Heimbuch et al(4) reported 1 J/cm^2^ dose may not be adequate to kill H1N1 influenza depending on the N95 respirator used. Mills et al found that only facepieces on 12 of 15 models and straps on 7 of 15 models showed a significant (≥ 3 log) reduction of H1N1 influenza viability. Similarly, Heimbuch et al found that only facepieces on 11 of 15 models and straps on 4 of 15 models showed a significant (≥ 3 log) reduction of H1N1 influenza viability.(8)

Some respirator models have materials, such as the straps of the 3M 1860, that demonstrate hydrophilic characteristics when inoculated. Moreover, these seemingly hydrophilic surfaces showed consistently lower mean log reduction < 3 log_10_ TCID_50_.(4) In contrast, seemingly hydrophobic materials, such as the 3M 1860 facepiece, were found to demonstrate a > 3 log_10_ TCID_50_ reduction.(4) In our study using SARS-CoV-2, we observed similar results with the 3M 1860 facepiece and strap. Further, the facepieces of the 3M 8210 have hydrophilic properties which were reflected in the reduced decontamination results, while the straps did not readily absorb the droplets, and hence were adequately disinfected. The Moldex 1511 facepiece and straps also appeared to be hydrophobic and did not absorb the droplets.

Some straps are prone to twisting. Consequently, when the respirator is flipped during the irradiation process, care must be taken to make sure the appropriate surface of the strap is exposed to UVC. Also, straps should not inadvertently lay on top of the respirator, hence creating a shadowing effect. Reduced decontamination seen amongst the straps may not only be a result of material but secondary to receiving a reduced dosage. UVC devices which provide 360 degrees of irradiation may obviate this issue. Possible respirator-based solutions include a secondary disinfection step (e.g. Environmental Protection Agency recommended cleansers) applied only to the straps. Further, ancillary disinfection testing was performed on the 3M 1860 straps using over-the-counter 70% isopropyl alcohol prep pads (TopCare, Elk Grove Village, IL). The straps were inoculated with SARS-CoV-2 and wiped three times with the alcohol pad. Results of the study showed that regardless of UVC irradiation, alcohol alone was sufficient to decontaminate the 3M 1860 straps (**Appendix Figure 2, Appendix Table 3**). Additionally, manufacturers may consider using, for example, the same material as the straps of the 3M 8210 for all the other models of FFRs to improve UVC decontamination.

Our dosage for this study was partially based on previous work with Influenza A (H1N1), Avian influenza A virus (H5N1), Influenza A (H7N9) A/Anhui/1/2013, Influenza A (H7N9) A/Shanghai/1/2013, MERS-CoV, and SARS-CoV,(4, 7-9) where it was determined that all areas of a respirator should receive at least 1 J/cm^2^. Preliminary unpublished data from the Photomedicine and Photobiology Unit at HFHS demonstrated through theoretical and measured models that the curvature and the distance of the 3M 1860 N95 respirator from the light source affected the dosage delivered in a predictable way. Moreover, extrapolating from this model, after irradiating one side of the respirator with 1.5 J/cm^2^, some of the lateral aspects may only receive 900 mJ/cm^2^ while the apex of the respirator may receive almost 3 J/cm^2^. Further, it was also observed that a certain percentage of the dosage received in an area (∼10%) permeates to the other side (I. Kohli, unpub. data). Therefore, 1.5 J/cm^2^ was chosen as the lowest irradiance to ensure that all areas received at least 1 J/cm^2^. Increasing the dosage delivered may improve decontamination, but UVC radiation can degrade certain polymers in a dose dependent manner, and the effects may vary greatly between different models.(10) Therefore, fit-testing of UVC decontaminated respirators must be performed each time a new model and/or dose is introduced into the healthcare system.(11)

Our study sampled different areas of each respirator to ensure that all ranges of dosages were accounted for in a real-world setting against SARS-CoV-2. Other strengths included the testing of different model types. Of note, the hydrophilic surfaces (e.g. 3M 8210 facepiece and 3M 1860 strap) of untreated positive controls demonstrated a lower virus recovery than control. Additional testing was performed to determine if the droplet was drying larger than the 4 mm area tested. The results showed that there was limited to no virus in the periphery of the 4 mm area tested, and that no virus could be detected on the wearer-facing surface. Moreover, the lower yield reflects a diminished ability to resuspend the virus after drying. Limitations of the study include that no soiling agents were used. However, at Henry Ford Health System, as in other healthcare facilities, personnel are instructed not to reuse respirators that are visibly soiled. Further, it is still unclear what the infectious dose is for SARS-CoV-2; therefore, it is unknown if a significant reduction in viral load eliminates contagious risk.

In conclusion, UVC at a dose of 1.5 J/cm^2^ applied to both sides is effective at decontaminating SARS-CoV-2 on some N95 respirators. This dose may only be an appropriate decontamination method to facilitate reuse of PPE for healthcare personnel when applied to certain models/materials. In addition, some straps may require additional disinfection to maximize the safety to the frontline workers. Implementation of widespread UVC decontamination methods requires a careful consideration of model, material type, design, and fit-testing following irradiation. It should also be emphasized that similar cautions should be practiced for all other methods of respirator decontamination.

## Data Availability

All data generated or analyzed during this study are included in this published article (and its supplementary information files).

---

**Appendix Table 1.** Preliminary testing of 3M 1860 N95 respirator. *SARS-CoV-2 = Severe Acute Respiratory Syndrome coronavirus 2; PBS = phosphate-buffered-saline; TCID_50_ = 50% tissue culture infectious dose; LOD = limit of detection.

| Log<sub>10</sub> TCID<sub>50</sub> / 4mm punch (300 µL) | Treated respirator A | Treated respirator B | Untreated respirator A | Untreated respirator B | 10 uL of SARS-CoV-2 stock | SARS-CoV2-stock |
| --- | --- | --- | --- | --- | --- | --- |
| Location 1 | < LOD | < LOD | 4.579181246 | 4.579181246 |  |  |
| Location 2 | < LOD | < LOD | 4.479181246 | 3.779181246 |  |  |
| Location 3 | < LOD | < LOD | 4.679181246 | 4.079181246 |  |  |
| Location 4 | 1.301029996 | < LOD | 2.479181246 | 2.379181246 |  |  |
| 10 µL of SARS-CoV-2 stock diluted in 300 uL of PBS |  |  |  |  | 4.679181246 |  |
| 25 µL of SARS-CoV-2 stock |  |  |  |  |  | 7.079181246 |
| Limit of detection | 1.301029996 |  |  |  |  |  |

**Appendix Table 2.**
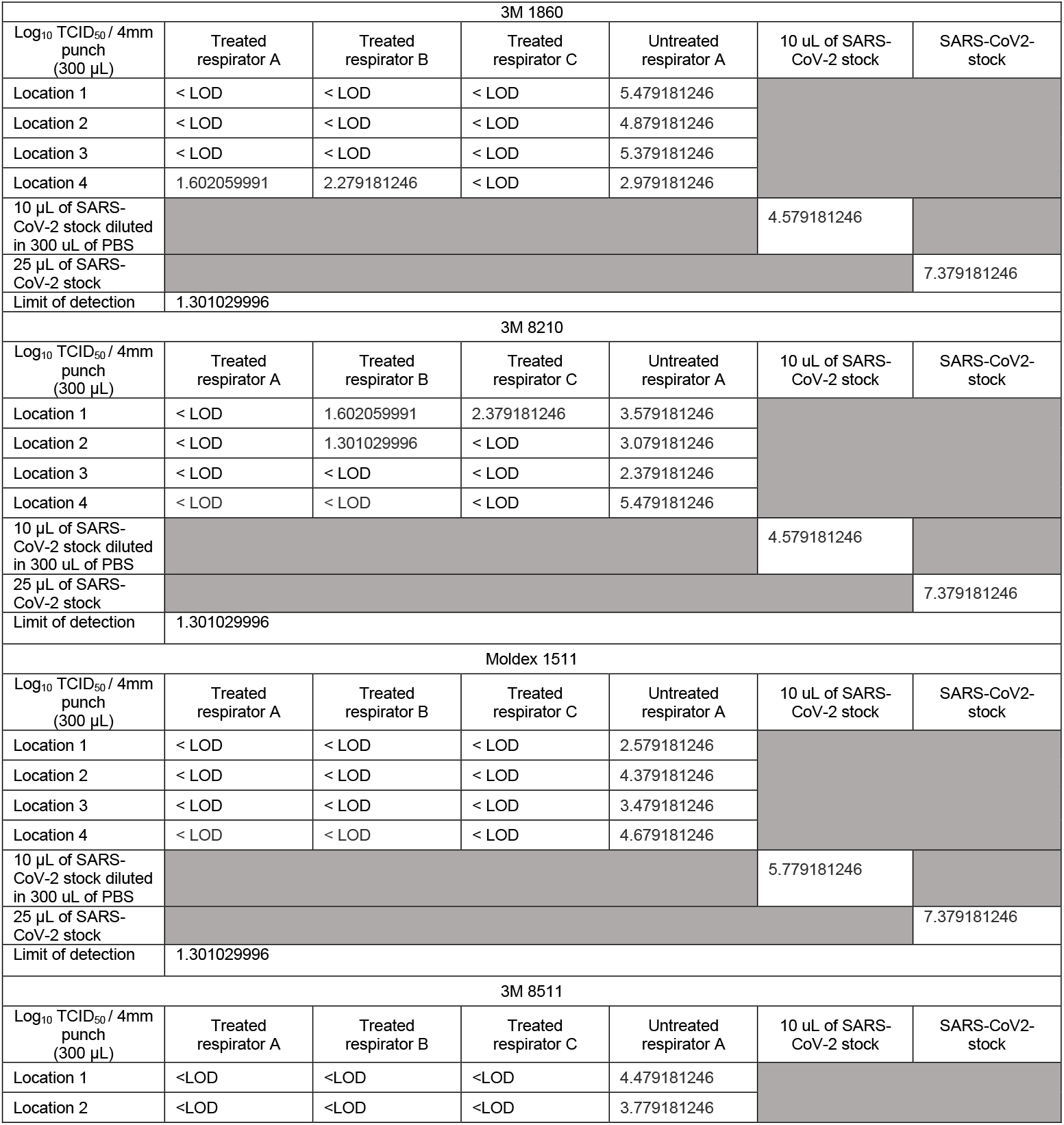
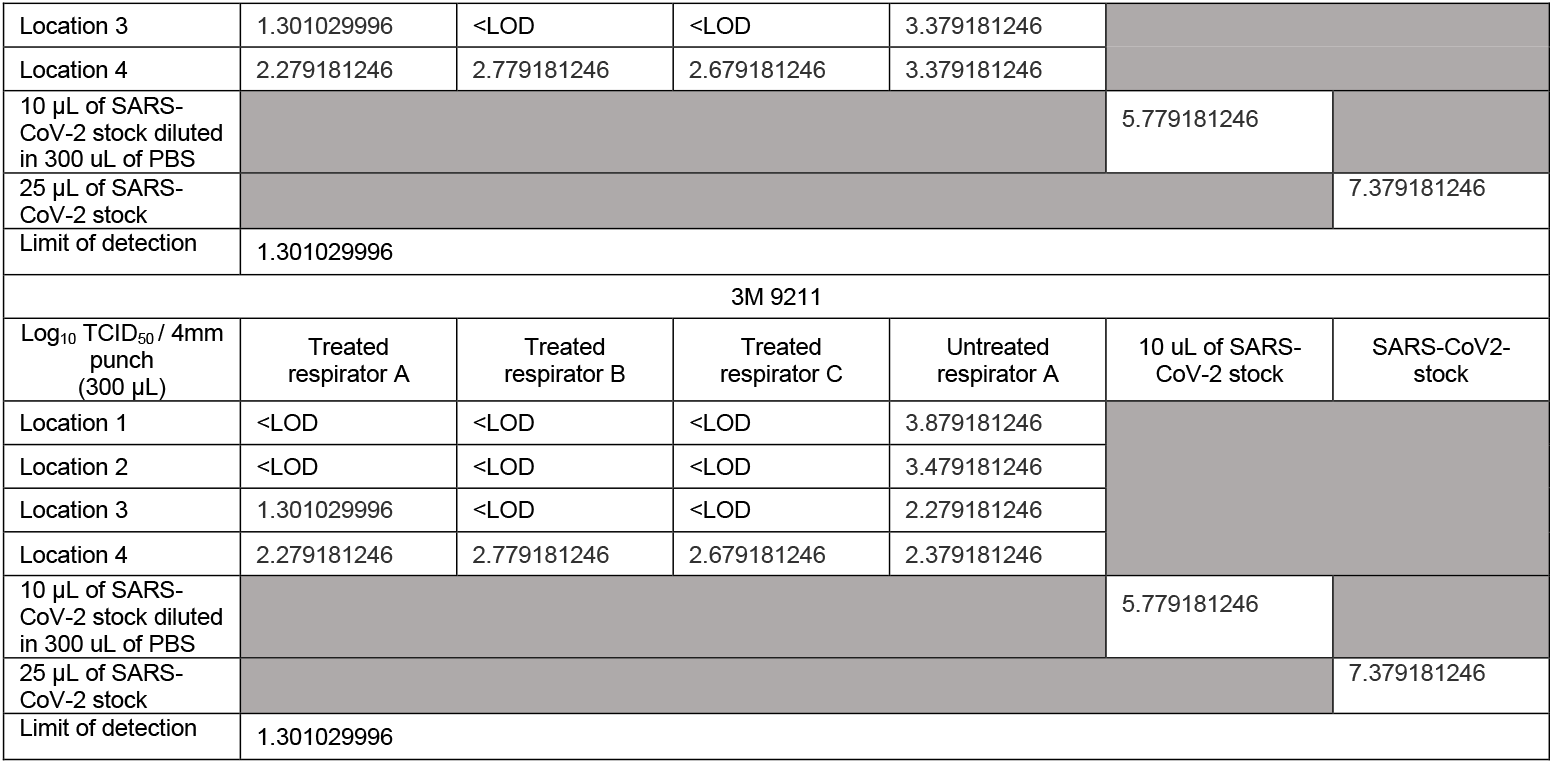
Recovered SARS-CoV-2 Log_10_ TCID_50_ / 4mm punch. *SARS-CoV-2 = Severe Acute Respiratory Syndrome coronavirus 2; PBS = phosphate-buffered-saline; TCID_50_ = 50% tissue culture infectious dose; LOD = limit of detection.

**Appendix Table 3.** Recovered SARS-CoV-2 Log_10_ TCID_50_ / 4mm punch on the 3M 1860 strap after secondary disinfection testing. *SARS-CoV-2 = Severe Acute Respiratory Syndrome coronavirus 2; PBS = phosphate-buffered-saline; TCID_50_ = 50% tissue culture infectious dose; LOD = limit of detection.

| 3M 1860 Strap |  |  |  |  |  |  |
| --- | --- | --- | --- | --- | --- | --- |
| Log<sub>10</sub> TCID<sub>50</sub> / 4mm punch (300 µL) | Alcohol Wiped Location 1 | Alcohol Wiped Location 2 | No Alcohol Wiped Location 1 | No Alcohol Wiped Location 2 | 10 uL of SARS-CoV-2 stock | SARS-CoV2-stock |
| UVC-Treated | < LOD | < LOD | 2.379181246047625 | 2.579181246047625 |  |  |
| UVC-Untreated | < LOD |  | 3.579181246047625 |  |  |  |
| 10 µL of SARS-CoV-2 stock diluted in 300 uL of PBS |  |  |  |  | 4.379181246047625 |  |
| 25 µL of SARS-CoV-2 stock |  |  |  |  |  | 5.779181246047625 |
| Limit of detection | 1.301029996 |  |  |  |  |  |

**Appendix Figure 1.**
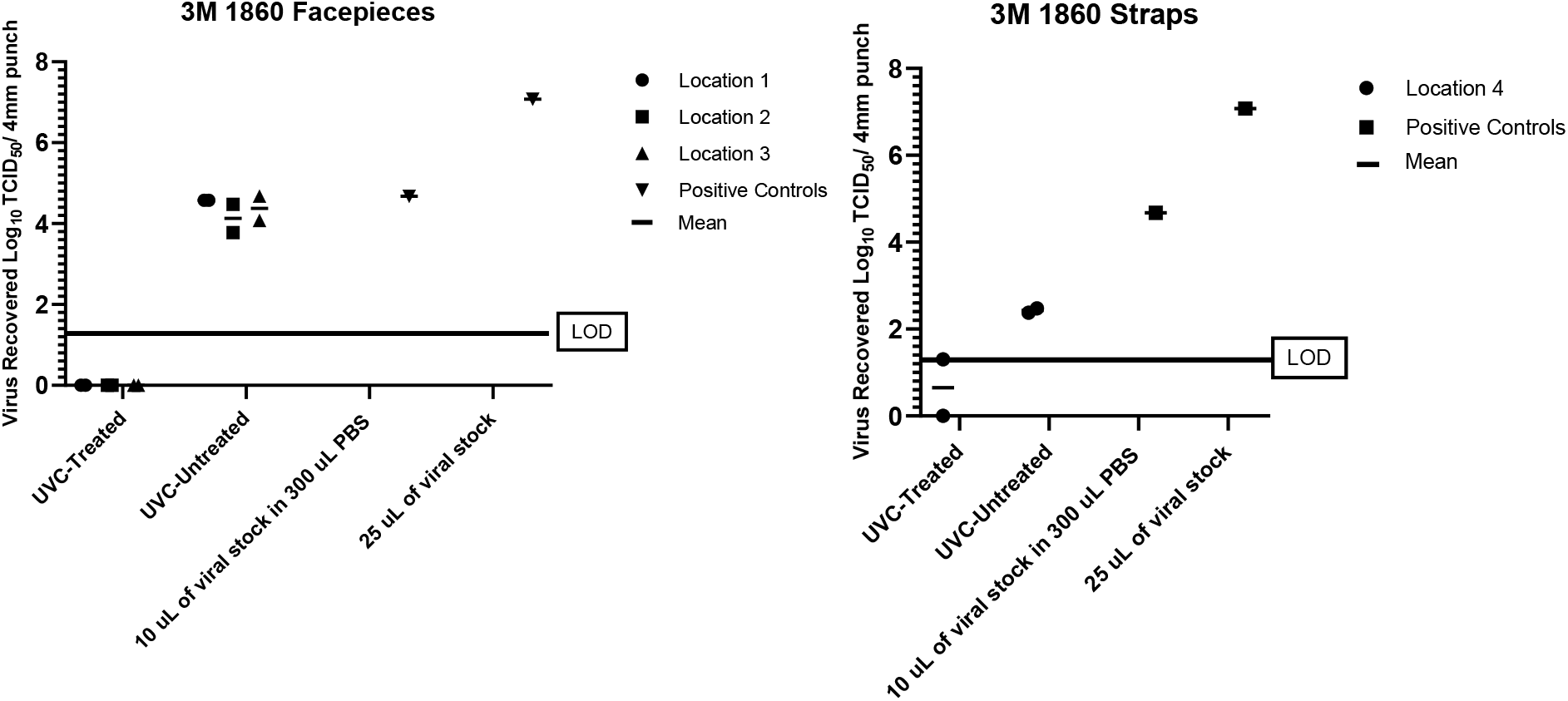
Preliminary testing of 3M 1860 N95 respirator. * Well that were below the limit of detection (LOD) and had no cytopathic effect were arbitrarily assigned the value of zero to represent this phenomenon in the above graphs. SARS-CoV-2 = Severe Acute Respiratory Syndrome coronavirus 2; PBS = phosphate-buffered-saline; TCID50 = 50% tissue culture infectious dose

**Appendix Figure 2.**
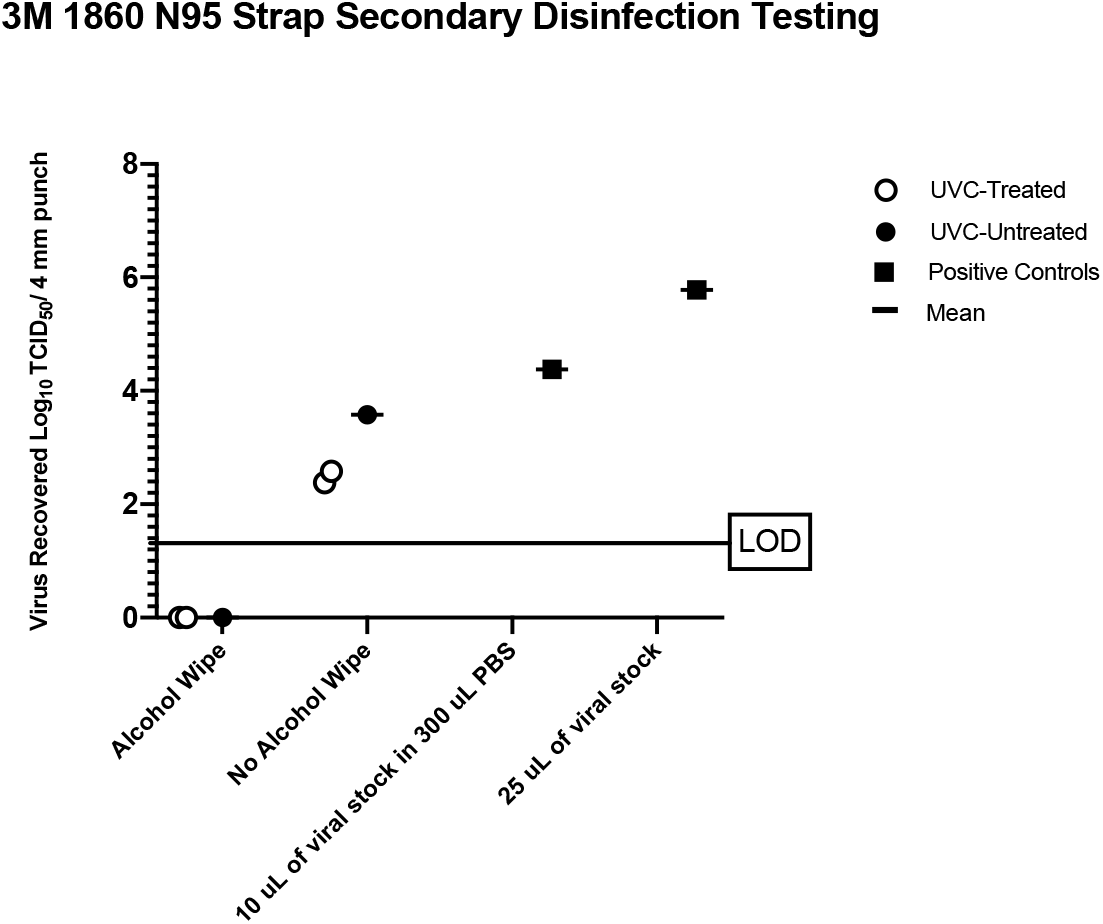
3M 1860 N95 Strap Secondary Disinfection Testing. *Wells that were below the limit of detection (LOD) and had no cytopathic effect were arbitrarily assigned the value of zero to represent this phenomenon in the above graphs. PBS = phosphate-buffered-saline; TCID50 = 50% tissue culture infectious dose; UVC = ultraviolet C.

